# Monocyte-to-Lymphocyte Ratio and Cardiovascular Outcomes in Individuals with Chronic Kidney Disease: A Systematic Review of Observational Studies

**DOI:** 10.1101/2025.10.14.25338013

**Authors:** Rostand Dimitri Messanga Bessala, Tchapleu Nana Ian Gidley, Ledoux Tresor Erlamala Pakourdey, Vuchichi Boris Vugugah, Richmond Nketia, Donnang Stanis, Jennifer Momo, Erna Hosanna Ngo Kam, Tasnim Sali, Jabo Jules, Carl Kamwa Vivian Njoya, Ngongang Ouankou Christian

## Abstract

**Introduction:** Inflammation contributes to increasing cardiovascular risk, especially in chronic conditions like chronic kidney disease (CKD). The monocyte-to-lymphocyte ratio (MLR) is a composite marker of inflammation. However, it remains relatively underexplored compared to other biomarkers. This study aimed to assess the MLR in relation to cardiovascular outcomes in CKD.

**Methods:** We systematically searched Medline, EMBASE, Web of Science, and Scopus from inception to May 28, 2025. We included peer-reviewed observational studies assessing MLR and cardiovascular outcomes or all-cause death in CKD. Two reviewers independently screened studies, extracted data, and assessed risk of bias using the Newcastle-Ottawa Scale. Findings were narratively synthesized.

**Results:** Eleven studies (n = 18,631) met our inclusion criteria. The study population ranged from non-dialysis CKD to end-stage kidney disease on dialysis, with follow-up from 1 to 124 months. Five studies (n = 16,974) examined cardiovascular death and generally reported significant associations with elevated MLR. Six studies (n = 4,587) assessed cardiovascular events except death, yielding inconsistent findings, although some reported significant associations. Five studies (n = 15,682) showed increased risk of all-cause death with increasing MLR. All except one of the eleven studies were rated as good quality.

**Conclusion:** Elevated MLR appears to be consistently associated with increased cardiovascular and all-cause death in CKD. Evidence for cardiovascular events remains inconsistent, and thresholds proposed in individual studies may not be generalizable. Large-scale, multi-ethnic, and prospective studies with standardized protocols are needed to validate MLR’s role in cardiovascular risk stratification.

## INTRODUCTION

### Description of the condition

Chronic kidney disease (CKD) refers to abnormalities of kidney structure or function that are present for at least 3 months and have implications for patient health. Diagnosis requires the presence of either one or more markers of kidney damage, including albuminuria (30 mg/g), urine sediment abnormalities, persistent hematuria, electrolyte imbalance or other abnormalities due to tubular disorders, histological changes, structural abnormalities identified by imaging, a history of kidney transplantation, or decreased glomerular filtration rate (GFR) < 60 ml/min per 1.73m^2^(^1^). Primarily caused by diabetes, CKD is an insidious, slowly progressive, and long-term condition that may remain asymptomatic until late stages(^2,3^). There are five stages of the disease: 1 (GFR ≥90 ml/min per 1.73m^2^), 2 (GFR = 89-60 ml/min per 1.73m^2^), 3a (GFR = 59-45 ml/min per 1.73m^2^), 3b (GFR = 44-30 ml/min per 1.73m^2^), 4 (GFR = 29-15 ml/min per 1.73m^2^), and 5 (GFR <15 ml/min per 1.73m^2^). Stage 5 is end-stage kidney disease (ESKD) and requires renal replacement therapy (RRT) in the form of dialysis or kidney transplantation(^1,4^). While CKD causes broad systemic complications, cardiovascular disease (CVD) remains the most significant (^2^), with a ten-to-twenty-fold higher risk compared to the general population(^5,6^). This burden highlights the need to further investigate the determinants of cardiovascular risk in CKD.

### Description of the exposure

Monocytes are white blood cells (WBC) that form part of the innate immune system and can differentiate into macrophages. Their role is key in the inflammatory response and they contribute to the pathogenesis of several chronic diseases, including atherosclerosis(^7^). In contrast, lymphocytes belong to the adaptive immune system and are responsible for regulating the immune response and maintaining immune memory(^8^). The monocyte-to-lymphocyte ratio (MLR), derived from circulating counts of these two immune cell populations, serves as a composite inflammatory biomarker that reflects the balance between innate and adaptive immune activity(^9^). An elevated MLR suggests heightened innate immune activation and a state of chronic low-grade inflammation. This is particularly relevant in the context of chronic diseases such as cancers, autoimmune conditions, and CKD. Given the well-established role of inflammation in the pathogenesis of CVD, inflammation biomarkers like MLR may offer valuable prognostic insights into cardiovascular risk among individuals with CKD(^10^).

### Why it is important to do this review

Immune dysregulation involving various leukocyte sub-populations, particularly neutrophils, monocytes, and lymphocytes, has been implicated in the inflammatory processes underpinning cardiovascular pathology in this group of patients(^11,12^). In this context, the MLR, a readily accessible and cost-effective inflammatory biomarker, has garnered growing interest. Several studies have explored the relationship between MLR and cardiovascular outcomes in CKD patients. However, the current body of literature is characterized by significant heterogeneity in terms of study design, patient populations, CKD stages, outcome definitions, and methods of MLR measurement and categorization. Consequently, findings have been inconsistent and difficult to interpret. This systematic review aims to synthesize the existing evidence of the association between MLR and cardiovascular outcomes in individuals with CKD, with the goal of providing a clearer understanding of its prognostic utility. (What does the MLR add that NLR/PLR do not?)

### Objectives

The main objective of this review was to systematically evaluate and synthesize the available evidence on the monocyte-to-lymphocyte ratio (MLR) and cardiovascular outcomes in individuals with CKD. Specifically, we aimed to:

i. assess the association between MLR and cardiovascular death in individuals with CKD;
ii. summarize the evidence regarding the association between MLR and cardiovascular events and all-cause mortality in individuals with CKD; and
iii. identify gaps in the current evidence and priorities for future research.

## METHODS

## PROTOCOL REGISTRATION

We used the Preferred Reporting Items for Systematic Reviews and Meta-analysis (PRISMA) guidelines to report our study findings, and no major changes were made with regard to the protocol. In addition, this systematic review was registered in the International Prospective Register of Systematic Reviews (PROSPERO) (CRD420251076059). Clinical trial number: not applicable

## ELIGIBILITY CRITERIA

### Criteria for considering studies for this review

#### Types of studies

Published peer-reviewed observational studies (cross-sectional and cohort studies) that explored MLR as a marker of cardiovascular risk in individuals with CKD were included. We limited our review to observational studies, as our preliminary scoping search revealed that existing literature on this topic comprised only observational designs. There were no language restrictions.

#### Types of participants

We included studies involving adult male and female patients aged *≥* 18 years old with diagnosed CKD (non-dialysis or dialysis-dependent) as defined by the KDIGO 2012: abnormalities of kidney structure or function, present for a minimum of 3 months, with implications for health, with either of the following criteria present(^1^):

- Markers of kidney damage (1 or more): Albuminuria (*ACR ≥ 30 mg/g [≥ 3 mg/mmol*]), urine sediment abnormalities, persistent hematuria, electrolyte and other abnormalities due to tubular disorders, abnormalities detected by histology, structural abnormalities detected by imaging, history of kidney transplantation;
- Decreased GFR: *GFR < 60 ml/min per 1.73m^2^*

We restricted our review to adults, as our preliminary search did not identify studies in children with CKD.

#### Types of exposure

The exposure considered in this systematic review is MLR obtained at baseline and/or measured repeatedly.

#### Types of outcome measures

In this systematic review, the focus was on cardiovascular outcomes. Cardiovascular death and events were the primary outcomes, and all-cause death was the secondary outcome.

Timing of outcome assessments.

Outcome measurement time points were extracted as described in the studies.

## INFORMATION SOURCES and SEARCH STRATEGY

### Search methods for identification of studies

#### Electronic searches

Four authors (R.D.M.B., R.N., C.K.V.N., and E.H.N.K.) developed the search strategies comprising subject headings and text words covering the concepts of MLR, cardiovascular risk, and CKD, which a fifth author (V.B.V.) reviewed (*Appendix 1*). We did not translate the included studies, as they were all published in English. To retrieve all possibly available studies, the database search started from inception. We searched the following databases until May 28, 2025:

- Medline via OVID (from inception in 1946 to present)
- Embase via OVID (from inception in 1947 to present)
- Web of Science Core Collection (from inception in 1964 to present)
- Scopus

#### Searching for other sources

We reviewed the reference lists of all retrieved articles on the same topic for other potentially relevant studies. While in the respective databases, we used the “related article” function to broaden the search. Only peer-reviewed sources were included in this systematic review.

## SELECTION and DATA COLLECTION PROCESS

### Data collection and analysis

#### Selection of studies

We imported search results into *Zotero version 7.0*(*13*) to remove duplicate records. Upon importing these results to the Rayyan AI(14), we manually removed further duplicates, and two review authors (R.D.M.B. and V.B.V.) independently screened the title and abstract of each record for potential relevance. We then retrieved full-text reports of the studies deemed relevant, and two review authors (R.D.M.B. and V.B.V.) independently screened them against the eligibility criteria to identify studies for inclusion. Any discrepancies that arose were settled by consensus with a third reviewer (C.K.V.N.). We recorded the study selection process in sufficient detail to complete a study selection flow diagram, a ‘Characteristics of included studies,’ and a ‘Characteristics of excluded studies.’

#### Data extraction and management

We developed a data extraction form that two review authors (R.D.M.B. and V.B.V.) piloted on five studies and adapted as required. Two review authors independently extracted data (R.D.M.B. and V.B.V.) and resolved discrepancies by consensus. We recorded the following information for each study:

- Bibliometric information: Names of authors, publication date, country
- Study characteristics: Study design, sample size, CKD stage, or dialysis status
- Participant characteristics: Age, gender
- Exposure characteristics: MLR values (cutoffs if any)
- Outcomes: Primary and secondary cardiovascular outcomes measured, effect sizes (hazard ratio, odds ratio and risk ratio, with corresponding 95% confidence intervals), and confounding variables adjusted for.

Assessment of risk of bias in included studies.

We performed the risk of bias assessment using the Newcastle-Ottawa Scale (NOS) for included studies. We selected the NOS as it is one of the most widely used tools for assessing the methodological quality of observational studies, particularly in systematic reviews. It covers key domains including participant selection, comparability, and outcome assessment, which align well with the designs of the studies included in our review(15). However, we acknowledge that the NOS could be subjective in scoring and may be limited in exploring biases. Two review authors (R.D.M.B. and V.B.V.) independently applied the risk of bias tool and resolved differences by discussion or appeal to a third review author (C.K.V.N.). Study quality was rated as good, fair, or poor.

#### Measures of Exposure Effect

The measures of exposure effect were cardiovascular death, cardiovascular events, and all-cause death.

#### Dealing with missing data

We only analyzed the available data, and there was no major adjustment or deviation from the registered protocol.

#### Data synthesis

Owing to substantial clinical and methodological heterogeneity across the included studies, such as differences in study populations, CKD stage, dialysis status, and effect estimates, a quantitative meta-analysis was not considered appropriate. Instead, findings were synthesized narratively. Descriptive statistics were used to summarize the characteristics of included studies. Where applicable, effect sizes and corresponding confidence intervals and p-values were extracted and tabulated according to outcome categories (cardiovascular death, cardiovascular events, and all-cause mortality). Frequencies and proportions were also used to categorize findings based on CKD stage and study design. Particular attention was given to the consistency and direction of the associations across studies, as well as differences in study design and patient populations. All visualizations were conducted using *R software version 4.5.0*(*16*).

## RESULTS

### Description of studies

#### Study selection (included studies)

Following a comprehensive search of the specified electronic databases and study reference lists, duplicates were removed, yielding a total of forty-two unique records. Of these, sixteen articles were retrieved for full-text screening. Ultimately, eleven studies met the predefined inclusion criteria. The study selection process is summarized in figure 1.

**Figure 1:**
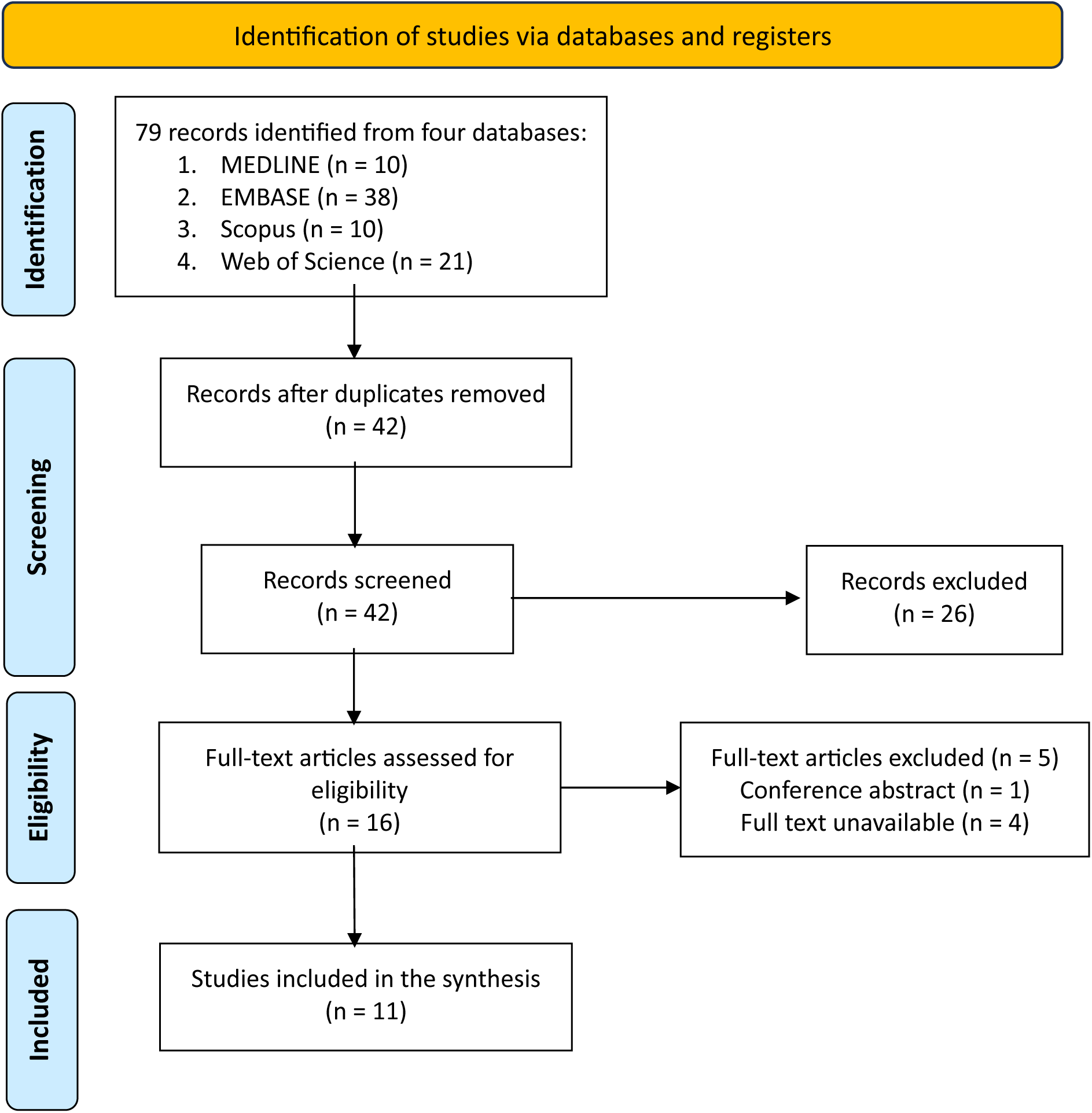
PRISMA flow chart describing the study selection process.

The selected studies collectively enrolled 18,631 participants. Study characteristics are presented in table 1. All included studies were published between 2018 and 2024. The study design comprised two cross-sectional(17,18), three prospective cohort studies(19–21), and six retrospective cohort studies(22–27). Follow-up duration varied considerably across studies. For instance, *Muresan et al.* reported the shortest follow-up period (one month), whereas *Liu et al.* and *Oh et al.* reported the longest, at 124 and 108 months, respectively(21,26,27). The remaining studies had follow-up periods ranging between 12 and 60 months (Table 1).

**Table 1:** Characteristics of observational studies evaluating the monocyte-to-lymphocyte ratio and cardiovascular outcomes in chronic kidney disease.

| Author | Country | Study design | n | Population | Follow-up<br>(months)* |
| --- | --- | --- | --- | --- | --- |
| *Han et al.* | China | Cross-sectional | 209 | ESKD | NA |
| *Wang et al.* | China | Cross-sectional | 387 | ESKD | NA |
| *Liu et al.* | USA | Retrospective cohort | 11262 | CKD | 124 |
| *Muto et al.* | Japan | Prospective cohort | 132 | ESKD | 49 |
| *Rezk et al.* | Egypt | Prospective cohort | 70 | ESKD | 12 |
| *Muresan et al.* | Romania | Retrospective cohort | 461 | ESKD | 1 |
| *Zhang et al.* | China | Retrospective cohort | 398 | ESKD | 60 |
| *Liao et al.* | China | Retrospective cohort | 213 | ESKD | 30 |
| *Oh et al.* | USA | Prospective cohort | 3391 | CKD | 108 |
| *Wen et al.* | China | Retrospective cohort | 1753 | ESKD | 31 |
| *Xiang et al.* | China | Retrospective cohort | 355 | ESKD | 51 |

Notes: \*Follow-up duration was rounded to the nearest whole number. *Han et al.* and *Wang et al.* conducted cross-sectional studies where patients are seen at one point in time (17,18). Consequently, follow-up does not apply to these studies; USA: United States of America; n: sample size; ESKD: end-stage kidney disease; CKD: chronic kidney disease; NA: not applicable

#### Study Selection (Excluded studies)

We excluded five studies following full-text review. One was excluded for being a conference abstract, while the remaining four were inaccessible in full texts despite institutional access.

#### Quality assessment of included studies

Quality assessment using the NOS is presented in table 2. Besides, all studies were rated as good quality except the study by *Muto et al.*(*20*), which was rated as fair.

**Table 2:** Quality appraisal of included observational studies based on Newcastle-Ottawa Scale.

| Study | Selection | Comparability | Outcome | Total | Quality |
| --- | --- | --- | --- | --- | --- |
| *Liu et al.* | 4 | 2 | 3 | 9 | Good |
| *Muto et al.* | 3 | 1 | 2 | 6 | Fair |
| *Rezk et al.* | 4 | 2 | 3 | 9 | Good |
| *Mureşan et al.* | 4 | 2 | 3 | 9 | Good |
| *Zhang et al.* | 3 | 2 | 3 | 8 | Good |
| *Liao et al.* | 4 | 1 | 2 | 7 | Good |
| *Oh et al.* | 3 | 2 | 3 | 8 | Good |
| *Wen et al.* | 4 | 1 | 3 | 8 | Good |
| *Xiang et al.* | 4 | 2 | 3 | 9 | Good |
| *Han et al.* | 3 | 2 | 3 | 8 | Good |
| *Wang et al.* | 4 | 2 | 3 | 9 | Good |

Notes: The figures represent the number of stars attributed to each domain of the Newcastle-Ottawa Scale

The principal methodological limitations identified across studies included inadequate adjustment for residual confounding, variability in participant selection, and differences in outcome ascertainment. Although most studies adjusted for major cardiovascular risk factors and CKD severity, adjustment for inflammatory conditions, medication use, and other potential confounders was inconsistent. These limitations should be considered when interpreting the reported associations between MLR and cardiovascular outcomes.

### Primary outcomes

#### Monocyte-to-lymphocyte ratio and cardiovascular death

Five cohort studies including a total of 16,974 participants investigated the association between MLR and cardiovascular death. MLR was categorized and analyzed variously: as quartiles(27), per doubling of MLR(21), as part of an inflammation score(24), and as tertiles(22,23). Overall, most cohort studies reported a positive association between elevated MLR and cardiovascular death, although the magnitude of the association varied across studies.

#### Effect estimates and corresponding confidence intervals have been rounded to the nearest two decimal places

Except for *Liao et al.* [HR (95% CI): 4.03 (0.88-18.38), p: 0.072] and *Liu et al.* [HR (95% CI): 2.17 (0.71-6.59), p: 0.093], all studies reported statistically significant hazard ratios (HR). To illustrate, *Liu et al.* and *Xiang et al.* found that elevated MLR in CKD participants was associated with a 117% and a 384% increase in the hazard of cardiovascular death, respectively(23,27). However, wide confidence intervals indicate imprecision around the estimates. *Oh et al.* observed a 27% higher rate of cardiovascular death, and *Wen et al.* reported a 45% increase associated with MLR (Figure 2).

**Figure 2:**
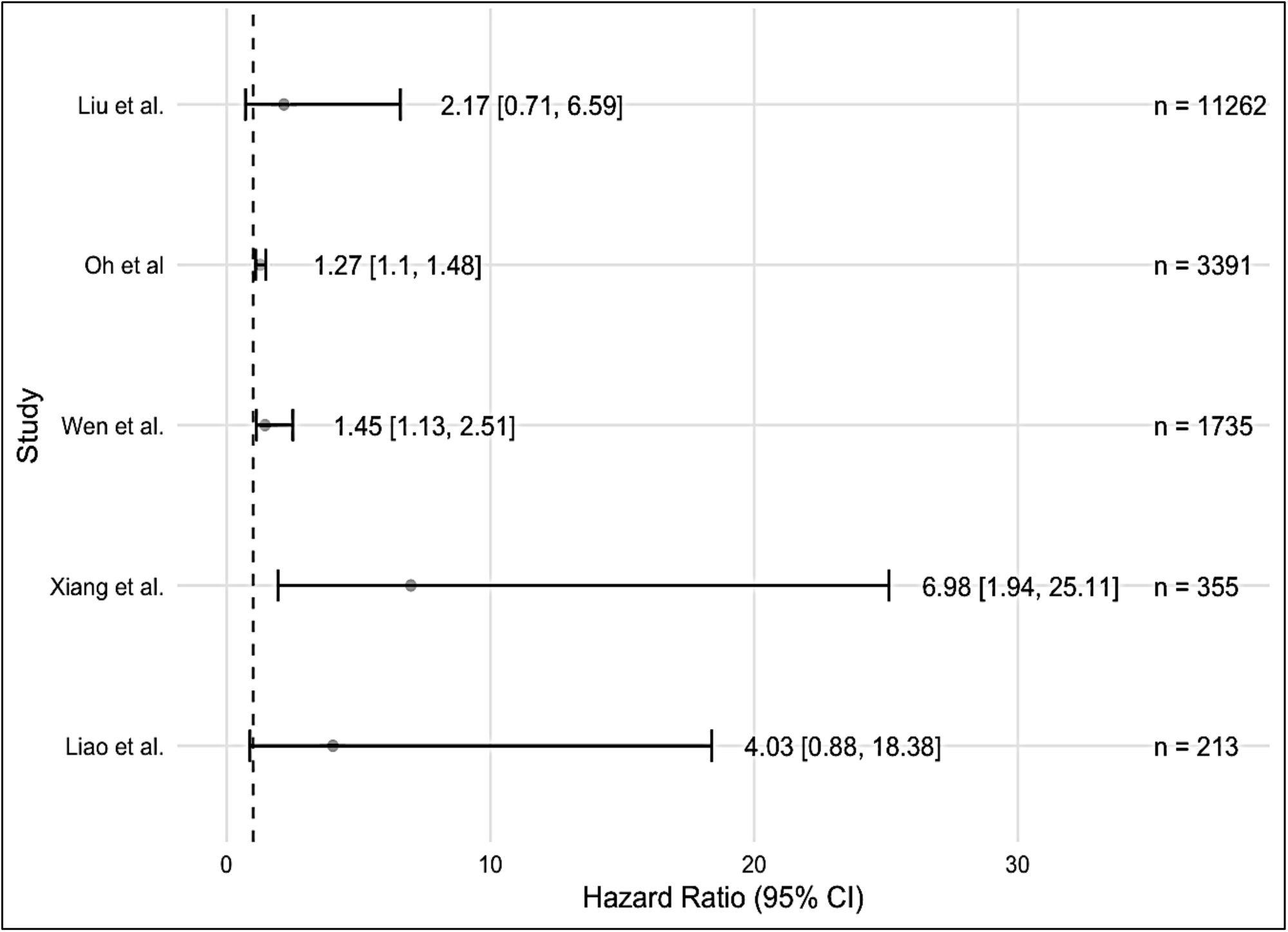
Summary of adjusted effect estimates of the association between monocyte-to-lymphocyte ratio and cardiovascular mortality in chronic kidney disease.

#### Monocyte-to-lymphocyte ratio and cardiovascular events

Six studies, comprising 4,587 participants, assessed the association between MLR and cardiovascular events, with inconsistent findings. *Muto et al.* reported that each unit increase in MLR was associated with a 5.56-fold increased risk of cardiovascular events in CKD(20) (Table 3). Similarly, *Oh et al.* observed that a doubling of MLR was associated with a 26% increase in cardiovascular event rates(21).

**Table 3:** Cardiovascular event outcome in relation to the monocyte-to-lymphocyte ratio in chronic kidney disease.

| Study | Cardiovascular outcome assessed | MLR comparison | Effect Type | Adjusted Estimate (95% CI) <sup>a</sup> | p-value <sup>b</sup> |
| --- | --- | --- | --- | --- | --- |
| *Han et al.* | Ischemic heart disease, Myocardial infarction, Heart failure, Atrial fibrillation, Valvular heart disease, Stroke (ischemic + hemorrhagic) | Inflammation index | OR | 2.51 (0.38-16.50) | 0.337 |
| *Muto et al.* | TIA, Angina, Peripheral Arterial Disease, Stroke, Sudden Death, Myocardial Infarction, Heart Failure | MLR change by 1 unit | RR | 5.56 (1.05-25.71) | 0.028 |
| *Rezk et al.* | TIA, Acute Coronary Syndrome, Unstable Angina, Ischemic Cardiomyopathy, Pulmonary Embolism, Biventricular Failure, Valvular Calcifications | MLR % change within 12 months | NA | - | 0.546 |
| *Oh et al.* | Heart Failure, Myocardial Infarction, Ischemic Stroke, Peripheral Arterial Disease | Per doubling of MLR | HR | 1.26 (1.16-1.36) | NR |

Notes: <sup>a</sup> Adjusted estimate for the most inclusive model. Effect estimates and corresponding confidence intervals have been rounded to the nearest two decimal places; <sup>b</sup> P-value for the adjusted estimate; Q: Quartile; MLR: monocyte-to-lymphocyte ratio; HR: hazard ratio; OR: odds ratio; RR: Risk ratio; CI: confidence interval; NR: not reported; NA: not applicable

Conversely, *Han et al.* and *Rezk et al.* reported no statistically significant association, suggesting that MLR may not consistently predict cardiovascular events in CKD(17,19) (Table 3). Further, *Zhang et al.* identified an MLR threshold of 0.43 as a significant predictor of cardiovascular events in patients with ESKD(25). In the same light, *Wang et al.* found that higher MLR tertiles—no suggested threshold—were significantly associated with the presence of carotid plaques, potentially indicating increased cardiovascular risk in CKD(18).

### Secondary outcome

#### Monocyte-to-lymphocyte ratio and all-cause death

The association between MLR and all-cause death was reported in five studies including overlapping populations and involving 15,682 participants(21,23,24,26,27). MLR was presented in different formats: quartiles(27), tertiles(23), per doubling(21), high versus low categories(26), as part of an inflammation score index(24). With the exception of *Muresan et al.*(26), who reported odds ratio (OR), all studies utilized HR as the primary effect measure.

#### Effect estimates and corresponding confidence intervals have been rounded to the nearest two decimal places

All studies reported statistically significant adjusted estimates, indicating a positive association between elevated MLR and all-cause death in CKD patients. For example, *Oh et al.* demonstrated that a doubling of MLR was associated with an 18% increase in all-cause death rate (HR: 1.18, 95% CI: 1.09-1.29)(21). *Muresan et al.* reported the highest effect estimate with an OR (95% confidence interval) of 9.46 (5.06-17.69)(26). Although statistically significant, most studies had wide confidence intervals, except for *Oh et al.,* indicating lower precision around the estimates (Table 4, Figure 3). Furthermore, *Muresan et al.* identified an optimal MLR cut off value of 0.63 for predicting all-cause death(26).

**Figure 3:**
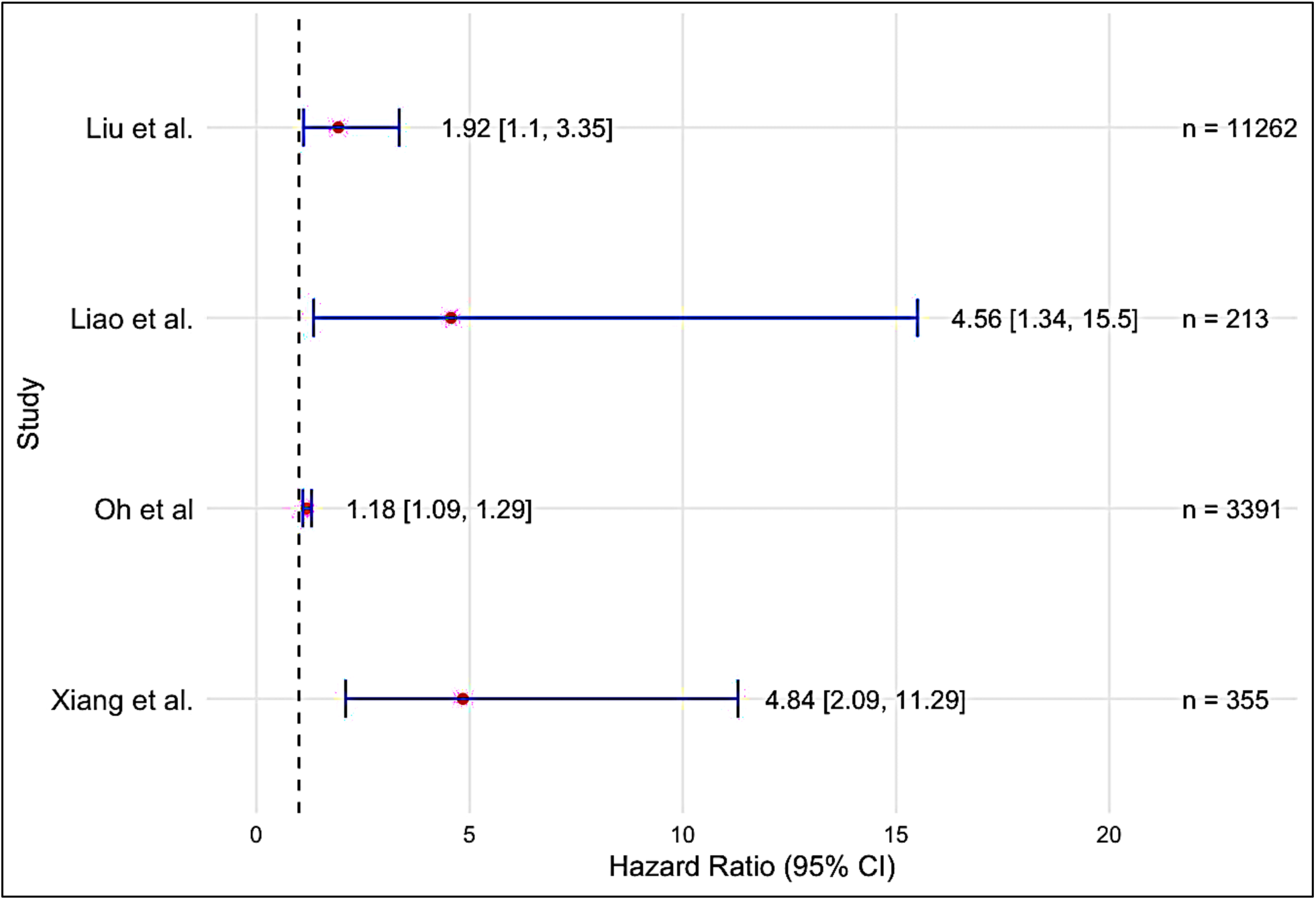
Summary of adjusted effect estimates of the association between monocyte-to-lymphocyte ratio and all-cause mortality in chronic kidney disease.

**Table 4:** Reported all-cause mortality and risk estimates based on MLR in included observational studies.

| Study | All-cause mortality outcome assessed | MLR comparison | Effect Type | Adjusted Estimate (95% CI)<sup>a</sup> | p-value<sup>b</sup> |
| --- | --- | --- | --- | --- | --- |
| *Liu et al.* | All-cause mortality | Q4 vs Q1 | HR | 1.92 (1.10-3.35) | 0.012 |
| *Muresan et al.* | All-cause 30-day mortality | High vs Low MLR | OR | 9.46 (5.06-17.69) | < 0.001 |
| *Liao et al.* | All-cause mortality | Inflammation score | HR | 4.56 (1.34-15.50) | 0.015 |
| *Oh et al.* | All-cause mortality | Per doubling of MLR | HR | 1.18 (1.09-1.29) | NR |
| *Xiang et al.* | All-cause mortality | MLR tertiles | HR | 4.84 (2.09- 11.21) | < 0.001 |

Notes: <sup>a</sup> Adjusted estimate for the most inclusive model. Effect estimates and corresponding confidence intervals have been rounded to the nearest two decimal places; <sup>b</sup> P-value for the adjusted estimate; Q: Quartile; MLR: monocyte-to-lymphocyte ratio; HR: hazard ratio; OR: odds ratio; CI: confidence interval; NR: not reported
The list of adjusted covariates is presented in appendix 2.

## DISCUSSION

In the past years, evidence pertaining to MLR as a biomarker of cardiovascular risk has grown. Unfortunately, this remains sparse, with only eleven observational studies meeting the inclusion criteria for this review. Concerning quality assessment, included studies were mostly assessed to be good. We observed a consistent positive association between MLR and cardiovascular death and all-cause death. This suggests that the MLR may increase cardiovascular and all-cause death in CKD patients. On the other hand, no decisive evidence was found for the association between MLR and cardiovascular events, as findings were conflicting. However, several limitations inherent to the studies and the review process must be critically considered when interpreting the findings.

First, the observational nature of all studies limits causal inference. Despite statistical adjustments for key confounding factors (*see appendix 2*), residual confounding from unmeasured variables remains a possibility. This methodological constraint weakens the internal validity of reported associations. Moreover, the MLR, while associated with inflammatory states, is a non-specific biomarker of systemic inflammation and can be influenced by a wide range of physiological and pathological processes such as infections, malignancies, and autoimmune conditions(28). The lack of standardization of MLR measurement, timing, and categorization (quartiles, tertiles, or continuous variables) further reduces cross-comparability of results. These methodological and biological limitations collectively undermine the reliability, reproducibility, and clinical translatability of the MLR as a prognostic marker in CKD, emphasizing the need for standardized approaches in future research.

Second, the possibility of reverse causation cannot be excluded. Given that most included studies were observational and relied on baseline MLR measurements, it remains unclear whether elevated MLR levels precede or result from underlying cardiovascular disease or systemic inflammation associated with CKD. Patients with subclinical cardiovascular disease may already exhibit elevated MLR due to chronic inflammatory activation, arguably creating the appearance of a predictive relationship. Without temporal sequencing or longitudinal and repeated MLR measurement, disentangling cause from consequence remains challenging.

Third, significant heterogeneity was observed across studies regarding population characteristics, CKD stage, dialysis status, follow-up duration, and outcome definitions. While some studies, such as that by *Oh et al*.(21), included only non-dialysis CKD patients, others, such as *Muto et al.* (20), focused specifically on patients undergoing hemodialysis or peritoneal dialysis, each with distinct cardiovascular risk profiles and inflammatory burdens. These might have likely influenced the magnitude and direction of associations. Additionally, the definitions and adjudication of cardiovascular events varied widely across studies. For instance, *Wang et al.* (18) relied on administrative coding and data, while *Oh et al.* (21) relied on clinical diagnoses. These discrepancies in outcome ascertainment introduce measurement bias and compromise the internal validity of pooled results. As a result, this heterogeneity not only precluded robust statistical pooling but also complicated interpretation of whether MLR truly performs consistently across all stages of CKD. This highlights a critical limitation in the evidence base: while our objective was to assess how MLR varies across CKD, the available data are too fragmented to establish a coherent pattern, particularly when dialysis status and stage-specific inflammatory burden are not accounted for.

Fourth, many studies provided limited information on serial MLR measurements, with limited consideration for temporal changes. While some studies relied on a single measurement of MLR at baseline, others provided insufficient details on the MLR measurement time intervals. For instance, *Han et al.*(17) assessed the MLR at a single point in time (at baseline). This approach assumes stability in the inflammatory status, which is unlikely in conditions like CKD. However, authors such as *Rezk et al.*(19) obtained repeated measures of the MLR. The absence of repeated measurements of MLR and sufficient details about measurement procedures and conditions undermines the differentiation between transient elevations due to acute conditions and sustained elevations due to chronic diseases. This distinction is critical since only a sustained increase in the MLR is likely to explain cardiovascular outcomes in CKD. Therefore, repeated measures of the MLR could allow for a more dynamic evaluation and risk stratification in CKD patients. Furthermore, while some authors provided predictive thresholds(25,26), the cut-offs were study-specific, derived from small populations, and variable. This undermines clinical utility and calls into question whether MLR can provide a standardized predictive threshold without rigorous, prospective validation. Our objective of identifying a meaningful cut-off could not be met with current evidence, exposing a critical gap in literature and the need for stronger evidence.

Lastly, the relatively small number and the geographical locations of the studies may have reduced the generalizability of our findings. While the number of studies was relatively small, the large sample size could possibly undermine the effect on the reliability of findings. In addition, most studies were conducted in Asian countries. This regional overrepresentation raises concerns about the applicability of findings to non-Asian populations, given existing differences in CKD progression, inflammatory profiles, and healthcare infrastructures and systems. In addressing our objective of identifying gaps to guide future research, it is evident that the literature is hampered by several shortcomings: a lack of diverse and multiethnic and multinational cohorts, minimal use of longitudinal MLR data, and inconsistent definitions of both exposure and outcomes. Unless these gaps are addressed, the role of MLR in predicting cardiovascular risk for CKD patients may remain speculative rather than evidence-based.

Although most included studies were rated as good quality using the Newcastle-Ottawa Scale, the overall body of evidence should be interpreted cautiously because all included studies were observational, heterogenous in their design, exposure definitions, outcome ascertainment, and adjustment strategies. Consequently, confidence in the overall findings remains limited, and prospective studies with standardized methodologies are needed to strengthen the evidence base.

Despite these limitations, this is the first systematic review, to the best of our knowledge, to comprehensively synthesize evidence on the association between MLR and cardiovascular risk in individuals with CKD. Further, the consistency of the association across studies and settings, including large national cohorts, strengthens the case for MLR as a potentially valuable and low-cost inflammatory biomarker, particularly in low-resource settings like sub-Saharan Africa, where access to high-sensitivity CRP or IL-6 testing is limited. As a result, MLR could provide a pragmatic tool for risk stratification. Future research should focus on prospective, multi-center studies that incorporate standardized MLR measurement, uniform cardiovascular outcome definitions, adjustment for dynamic changes over time, and time-to-event analyses. Such studies would help to ascertain the clinical utility of MLR in risk prediction models, clarify the direction of association, and elucidate its potential role in guiding targeted interventions to reduce cardiovascular burden in CKD.

### Conclusions

In conclusion, this systematic review highlights a growing but still limited body of evidence supporting the MLR as a prognostic biomarker for cardiovascular death and events, as well as all-cause death in individuals with CKD. MLR appears promising but is not yet ready for incorporation into routine cardiovascular risk prediction models. Future large-scale, multiethnic, and prospective studies with standardized protocols can help in determining whether the MLR has a legitimate role in improving cardiovascular risk prediction, thus guiding therapeutic interventions in CKD care.

## DECLARATIONS

### Ethical approval

No ethical approval was required for this systematic review, as authors worked exclusively with published studies.

### Availability of data and materials

All datasets used and/or analyzed during the current study are available from the corresponding author on reasonable request.

### Authors’ contributions

Authorship for this systematic review was based on the International Committee of Medical Journal Editors (ICMJE) and participants to this review who did not fulfil all of the four criteria were considered as non-author contributors and acknowledged in the final manuscript.

*Project conceptualization: R.D.M.B., V.B.V., and N.O.C. Search strategy: R.D.M.B., R.N., C.K.V.N., and E.H.N.K. Data analysis and synthesis strategy: R.D.M.B. and R.N. Protocol writing: R.D.M.B. and V.B.V. Protocol revision: R.D.M.B. Data Screening: R.D.M.B. and V.B.V. Data extraction: R.D.M.B., and V.B.V. Study quality assessment: R.D.M.B. and V.B.V. Data analysis and synthesis: R.D.M.B., T.N.I.G., L.T.E.P., T.S., and R.N. Manuscript drafting and editing: R.D.M.B., V.B.V., R.N., C.K.V.N., T.N.I.G., L.T.E.P., D.S., J.M., T.S., J.J., E.H.N.K, and N.O.C*.

## Data Availability

The extracted dataset supporting the findings of this study will be made publicly available on OSF or Zenodo upon publication of the preprint and once available.

https://osf.io

https://zenodo.org

## Acknowledgments

This systematic review did not contribute towards a degree award. We would like to thank Dr SOH MANKONG Dorothee for her critical and timely comments on our project.

## APPENDICES

### Appendix 1: Search strategy (search terms and string from inception until May 28, 2025)

(“monocyte lymphocyte ratio” OR “monocyte to lymphocyte ratio” OR “MLR”) AND (“chronic kidney disease” OR “CKD” OR “renal insufficiency” OR “end-stage renal disease” OR “ESRD”) AND (“cardiovascular disease” OR “CVD” OR “cardiovascular risk” OR “cardiovascular events” OR “cardiovascular mortality” OR “myocardial infarction” OR “stroke”).

### Appendix 2: List of adjusted covariates

| Author | Adjusted covariates for the most inclusive model |
| --- | --- |
| *Liu et al.* | Age, sex, race, education level, smoking status, hypertension, diabetes, CVD, history of ACEi and statin, BMI, SBP, total cholesterol, HbA1c, BUN, eGFR, CRP, albumin |
| *Muresan et al.* | Atrial fibrillation, myocardial infarction, chronic heart failure, diabetes, peripheral artery disease, smoking status, dialysis access type, inflammatory biomarkers (NLR, MLR, PLR) |
| *Liao et al.* | Age, sex, BMI, primary cause of ESKD, MAP, 24h urine output, hemoglobin, calcium, phosphorus, albumin, iPTH, total cholesterol, Triglycerides, |
| *Muto et al.* | Age, sex, diabetes mellitus, smoking status, CRP |
| *Han et al.* | Age, sex, BMI, dialysis time, smoking, alcohol consumption, primary diseases, HADS-A score, HADS-D score, DUREL, Serum creatinine, glucose, albumin, PAB, Magnesium |
| *Oh et al.* | Age, sex, race, clinic site, traditional CVD risk factors (systolic blood pressure, number of antihypertensives, total cholesterol, HDL cholesterol, statin use, smoking status, prevalent CVD, and prevalent diabetes), markers of kidney disease (eGFR and urinary albumin), hs-CRP |
| *Xiang et al.* | Age, sex, time on HD, history of CVD, diabetes mellitus, hypertension, BMI, TG, LDL, HDL, cholesterol, albumin, hemoglobin, smoking status, calcium, phosphate, iPTH, ferritin, White cell count, neutrophil count, lymphocyte count, monocyte count, hs-CRP, MLR, NLR |
| *Wen et al.* | Age, sex, CCI, current smoking, current drinking, ACEi/ARB use, Beta blocker use, statin use, serum albumin, total cholesterol, Triglycerides, LDL, HDL, serum uric acid, hs-CRP, NT-pro BNP, eGFR |
| *Wang et al.* | Age, sex, BMI, eGFR, smoking status, diabetes mellitus, hypertension, hemoglobin, serum albumin, cerebrovascular and cardiovascular disease |

### Appendix 3: Abbreviations

ACR: Albumin-Creatinine ratio; ACEi: Angiotensin converting enzyme inhibitors; AI: Artificial intelligence; ARB: Angiotensin 2 receptor blocker; BMI: Body mass index; BUN: Blood urea nitrogen; CVD: Cardiovascular disease; CCI: Charlson comorbidity index; CKD: Chronic kidney disease; DUREL: Duke University Religion index; ESKD: End-stage kidney disease; eGFR: Estimated glomerular filtration rate; HADS: Hospital Anxiety Depression Scale; HDL: High-density lipoprotein; hs-CRP: High sensitivity C reactive protein; iPTH: Intact parathyroid hormone; KDIGO: Kidney Disease Improving Global Outcomes; LDL: Low-density lipoprotein; MAP: Mean arterial pressure; MLR: Monocyte-to-Lymphocyte ratio; NLR: Neutrophil-to-Lymphocyte ratio; NOS: Newcastle-Ottawa Scale; NT-pro BNP: N-terminal pro type brain natriuretic peptide; PAB: Prealbumin; PLR: Platelet-to-lymphocyte ratio.

